# Auditory-induced Affective State Modulates Left DLPFC Engagement in Response to iTBS: An fNIRS Study in Healthy People

**DOI:** 10.64898/2026.11.26.26369021

**Authors:** Nancy M.X.Y. Shi, Alvin H.P. Tang, Minxia Jin, Adam W.L. Xia, Rebecca L.D. Kan, Bella B.B. Zhang, Penny P. Qin, Tim T.Z. Lin, Sharie X. Wang, Vera W.M. Lam, Wanda M.W. Chau, Frank Padberg, Juha Silvanto, Georg S. Kranz

## Abstract

**Background:** Intermittent theta burst stimulation (iTBS) of the left dorsolateral prefrontal cortex (DLPFC) is widely used as antidepressant treatment, yet response variability remains substantial. Cortical plasticity is state-dependent, and the affective state is a contributing factor. As the left DLPFC is both the iTBS target and a key node in affect regulation, we used fNIRS to measure its hemodynamic responses to stimulation during affective modulation.

**Method:** Thirty-six healthy participants received iTBS of the left DLPFC at 70% resting motor threshold intensity during positive- and negative-valence auditory stimuli compared to neutral-valence auditory stimuli as a control, with concurrent fNIRS recording across baseline, during-stimulation, and post-stimulation phases. Affective induction was confirmed via Self-Assessment Manikin ratings.

**Results:** Affective induction was successful. A two-way repeated-measures ANOVA revealed significantly lower oxygenated hemoglobin (HbO) in the affective state relative to control (main effect of State; F_1,35_ = 6.52, p = 0.015). For deoxygenated hemoglobin (HbR), a significant State × Time Phase interaction emerged (F_1,35_ = 6.00, p = 0.019): HbR was significantly higher during stimulation than post-stimulation in the affective state (p < 0.001), but not in the control state. Linear mixed-effects models comparing 20 time windows (w_1_-w_20_, each averaged 20 seconds from audio onset) against baseline showed no significant HbO changes but distinct HbR temporal dynamics.

**Conclusion:** Auditory-induced affective state modulates left DLPFC engagement in response to iTBS, with overall state differences reflected in HbO, and fine-grained temporal dynamics captured by HbR. These findings highlight the potential value of considering affective state in neuromodulation strategies.

## 1. Introduction

Non-invasive brain stimulation (NIBS) techniques are valuable research tools for modulating neural activity. Because of their high tolerability and non-surgical nature, NIBS techniques have generated widespread clinical interest for therapeutic applications in various neurological and psychiatric disorders^1^. Among NIBS modalities, transcranial magnetic stimulation (TMS) delivers transient electromagnetic fields to target brain regions, with repetitive applications (rTMS) enhancing neuroplasticity^2^. In particular, intermittent theta-burst stimulation (iTBS) is an advanced excitatory form of rTMS, reducing session times to approximately 3 minutes while maintaining comparable therapeutic efficacy^3–5^.

The dorsolateral prefrontal cortex (DLPFC) plays a crucial role in diverse psychological processes, including cognitive control^6^, working memory^7^, attention^8^, and affect regulation^9^. In many countries, iTBS of the left DLPFC is an approved treatment for patients with major depressive disorder (MDD)^10^. However, substantial inter- and intra-individual variability in treatment response remains a major challenge for clinical outcomes^11,12^. A leading explanation for these differences involves the concept of state-dependency in cortical plasticity: a fundamental property of neural processing wherein the effects of external stimulation depend not only on applied stimulation parameters, but also on the functional state of the targeted neural population^11,13^. Key state-dependent factors on the neural level include spontaneous oscillatory fluctuations and prior stimulation history, and ongoing cognitive or affective states on the psychological level^14,15^. While recent efforts have sought to optimize therapeutic TMS efficacy by leveraging real-time neural oscillations for closed-loop stimulation, employing priming stimulation protocols, or integrating concurrent cognitive tasks^16^, the influence of affective states has received limited attention. How affective states modulate iTBS target engagement, along with the underlying neural mechanisms, remains largely unexplored.

Valence is a fundamental dimension of affective space, spanning positive (pleasant) and negative (unpleasant) polarities. Its neural processing engages a shared brain network^17^ exhibiting a U-shaped activation pattern^18^, with both affective extremes eliciting heightened responses relative to the neutral condition. A recent study demonstrated that DLPFC activation during autobiographical memory retrieval exhibits a significant interaction between psychological resilience and valence, prompting the concept of affect-contingent neuromodulation as a potential strategy^19^. Given the dual role of the DLPFC as both a primary stimulation target and a critical region for affect regulation^20,21^, we aimed to investigate whether affective states influence the hemodynamic responses to left DLPFC-targeted iTBS.

Functional near-infrared spectroscopy (fNIRS) is a safe, portable, and reliable optical neuroimaging technique that monitors functional brain activity by leveraging neurovascular coupling (NVC) to detect differential near-infrared light absorption by oxygenated (HbO) and deoxygenated (HbR) hemoglobin^22^. Because fNIRS-derived hemodynamic responses correlate strongly with blood-oxygen-level-dependent (BOLD) fMRI signals, the gold standard for functional neuroimaging^23–25^, fNIRS is widely used to assist in the clinical assessment of brain dysfunction and in basic neuroscience research across multiple pathologies^26,27^. Importantly, its immunity to electromagnetic interference makes it uniquely suited for concurrent pairing with rTMS. We therefore employed concurrent fNIRS to record hemodynamic responses to iTBS in the left DLPFC across different states induced by auditory stimuli. We primarily hypothesized that iTBS-evoked hemodynamic responses would differ significantly between affective and control states. Our secondary objective was to analyze the detailed temporal trajectories in each state to identify the time-course features and trends underlying these differences.

## 2. Methods

### 2.1 Participants

This study was approved by the Institutional Review Board of the Hong Kong Polytechnic University (HSEARS20230324002) and pre-registered at ClinicalTrials.gov (NCT06750614), with the current report focusing specifically on the affective states. Participants were compensated for their time at a rate of 150 HKD (∼19.15 USD) per hour.

Thirty-six healthy participants, who were right-handed (self-report), were recruited via on-campus advertisements from December 2024 to December 2025. Respondents were screened by phone in chronological order of registration. A 1:1 sex ratio was maintained to ensure balanced representation, given the influence of sex differences on cortical excitability ^28,29^. Eligible individuals were scheduled for an in-person visit, where they received a comprehensive explanation of the study procedures and provided written informed consent in accordance with the Declaration of Helsinki. Subsequent assessments included a health status evaluation, a TMS safety screening questionnaire^30^, and demographic information collection.

The inclusion criteria were: (1) age between 18 and 60 years; (2) right-handedness, assessed using the Chinese version of the Edinburgh Handedness Questionnaire^31^; and (3) a Patient Health Questionnaire (PHQ-9)^32^ score ≤ 4. The exclusion criteria were: (1) a history of any psychiatric or neurological disorders; (2) any active or chronic medical conditions; (3) a first-degree relative diagnosed with a psychiatric disorder; and (4) a background/education in psychology or neuroscience and related fields.

### 2.2 Study design

#### Procedure

We employed a randomized, crossover design. Participants attended three experimental sessions, each incorporating a specific condition (Neutral, Positive, or Negative), presented in a randomized order and separated by a washout period of at least 3 days to prevent potential carryover effects^33^. Concurrent fNIRS was used to monitor real-time cortical hemodynamic responses. Once setup was complete, participants lay still with their eyes closed in a quiet, dimly lit environment during the experimental process. The experiment started with a 5-minute resting acclimatization period, the final 30 s of which served as the baseline. Following verbal instructions, the assigned condition commenced 8 s before iTBS onset and continued throughout the stimulation period. Each session concluded with a 200 s post-stimulation resting period, after which participants reported any discomfort and rated their pain level during stimulation using a Visual Analog Scale (VAS). To minimize potential confounding factors affecting brain activity^34^, all sessions were scheduled in the afternoon, and participants refrained from caffeine, alcohol, and strenuous physical exercise for 24 hours before each visit. The experimental procedure and setup are illustrated in Figure 1a and b.

**Figure 1.**
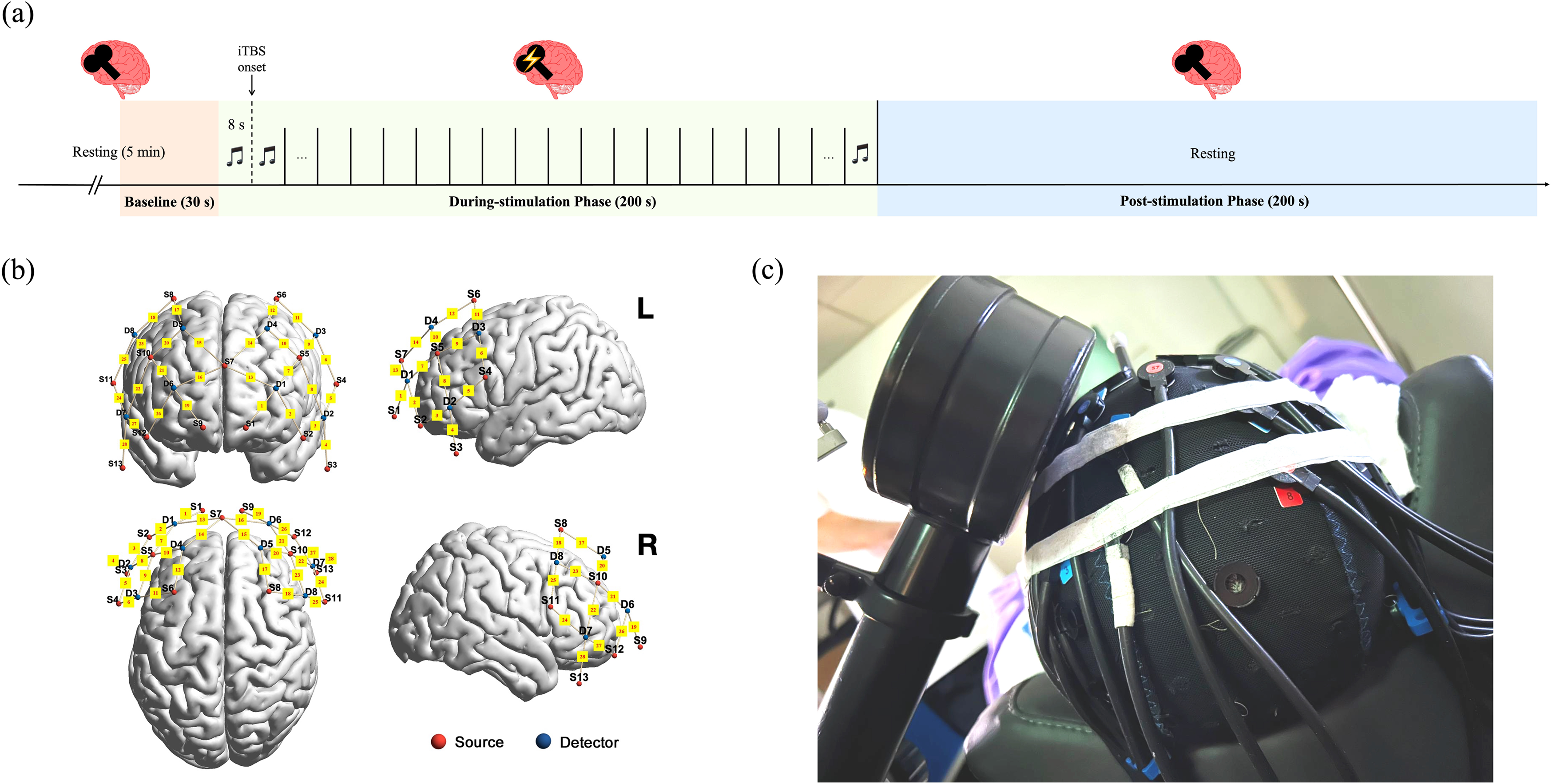
Experimental design and setup. **(a)** Diagram of the experimental procedure; **(b)** Concurrent iTBS/fNIRS setup; **(c)** fNIRS standard channel locations.

#### States and conditions

The two states were: (1) an affective state (comprised of Positive and Negative conditions), which manipulated affect through acoustic stimuli with an distinct affective valence; and (2) a control state (Neutral condition), which controlled for auditory input without affective induction. Acoustic stimuli were selected from the Expanded Version of the International Affective Digitized Sounds (IADS-E database)^35^, which contains 935 standardized 6-second audio clips covering a diverse range of daily life sounds. These clips are characterized by ratings on the 9-point Self-Assessment Manikin (SAM) scale^35,36^. Based on their valence scores (1 = most unhappy; 9 = most happy), 25 audio clips were selected: the 10 highest-valence positive audio clips, 10 valence-matched negative audio clips, and 5 neutral audio clips (mean valence = 5.00).

To implement an interleaved iTBS and auditory stimulation protocol, the selected audio clips were compiled into audio tracks with sound played exclusively during the 8-second inter-train intervals. Each clip was looped to fill the window and presented in a fixed pseudo-random sequence, consisting of two cycles for the Positive and Negative conditions (10 clips × 2) and four repetitions for the Neutral condition (5 clips × 4). In all conditions, audio tracks were preceded by the verbal cue “Ready, start”. Following each session, subjective SAM ratings of participants’ affective experiences during iTBS were collected to confirm successful induction.

#### TMS and Neuronavigation

TMS pulses were delivered using a MagPro X100 magnetic stimulator equipped with an active cooling figure-of-eight coil (Cool-B65; MagVenture A/S, Farum, Denmark). During the first visit, the left M1 hand area was initially located using standard Montreal Neurological Institute (MNI) coordinates (x = -42, y = -16, z = +68; Brodmann area 4, BA4) guided by a neuronavigation system (LOCALITE, Bonn, Germany), followed by manual optimization to identify the exact individual motor hotspot. To account for the increased coil-to-cortex distance introduced by the concurrent setup, we determined the resting motor threshold (RMT) while participants wore the fNIRS cap. RMT was defined as the minimum stimulator output intensity required to elicit motor evoked potentials (MEPs) with a peak-to-peak amplitude ≥ 50 μV in at least five out of ten consecutive trials from the contralateral first dorsal interosseous (FDI) muscle at rest.

During each session, standard iTBS was applied over the left DLPFC (x = -38, y = +44, z = +26; Brodmann area 46, BA46) at 70% of the individual RMT. This intensity was chosen because it provides efficacy comparable to the original 80% active motor threshold (AMT) iTBS protocol developed by Huang et al.^37,38^ and exerts considerable excitatory effects on neural activity^39^, while being better tolerated and producing less muscle twitching that could interfere with fNIRS signal quality. Otherwise, the current parameters followed standard iTBS protocols (i.e. 600 pulses per session, 2 s train of 10 bursts, with 3 pulses at 50 Hz, and 200 ms inter-burst interval, delivered every 10 s for a total of 20 trains)^37^.

#### fNIRS and regions of interest (ROI)

Hemodynamic signals were recorded at a sampling rate of 6.26 Hz using a dual-wavelength (785 and 830 nm) continuous-wave fNIRS system (NIRx Borealis; NIRx Medical Technologies, Berlin, Germany). Positioned according to the international 10-20 EEG system, the optode montage covered the bilateral DLPFC, comprising 13 sources and 8 detectors with a standard source-detector distance of 30 mm. In addition, 8 short-separation channels (SSC) were integrated into a coupler to record superficial scalp blood flow.

Identically configured fNIRS caps were available in three sizes (54, 56, and 58 cm) and fitted to individual head circumference. Optode localization and channel selection followed established protocols^29,40^. The stimulated DLPFC (i.e., the left DLPFC) was defined as the region of interest (ROI), corresponding to Ch-7 for the 56 cm cap and Ch-2 for the 54 and 58 cm caps. The optode standard channel distribution is shown in Figure 1c.

### 2.3 fNIRS data processing

Raw fNIRS signals were preprocessed using the Homer2 toolbox^41^ and custom MATLAB scripts (version R2024b; The MathWorks, Natick, MA, United States). Processing steps mainly followed the established analytical frameworks^29,40^. First, channel signal quality was checked via spectral analysis^42^, and channels lacking a distinct cardiac peak were rejected. Data were excluded if the target ROI channel or more than 50% of all channels exhibited poor signal quality. Raw signals from the remaining data were converted into optical density, and relative changes in hemoglobin concentrations (ΔHbO and ΔHbR) were calculated using the modified Beer–Lambert law^43^.

Subsequently, the following analyses were performed independently for ΔHbO and ΔHbR: (1) Short-separation regression (SSR): to eliminate superficial scalp hemodynamics, the most correlated SSC was identified for each standard channel and regressed out^44^. (2) Principal component analysis (PCA): to reduce signal dimensionality, PCA was performed to retain the principal components (PCs) explaining at least 99% of the total data variance^45^. (3) Independent component analysis (ICA): to mitigate deep breathing and motion artifacts, ICA was conducted with the number of independent components (ICs) set equal to the number of retained PCs. ICs displaying U-shaped or inverted-U-shaped patterns^46^, or multiple transient peaks synchronized with the iTBS pulses were discarded. (4) Wavelet transformation: to denoise residual artifacts, wavelet-based filtering was applied using a threshold of α = 0.1^47^. (5) Timeline reconstruction: to standardize the signal duration across all recordings, the time series were trimmed to include a 30-second pre-stimulation baseline, a 200-second concurrent task/iTBS during-stimulation phase, and a 200-second post-stimulation phase. (6) Band-pass filtering: to isolate stimulation-evoked hemodynamic responses, the physiological signals (heartbeat ∼1 Hz, respiration ∼0.3 Hz, Mayer waves ∼0.1 Hz, neurogenic regulations ∼0.04 Hz^48^) and low-frequency signal drift (1/430s ≈ 0.0023 Hz) were filtered using cut-off frequencies of 0.0023 – 0.04 Hz. (7) Baseline correction: to ensure that iTBS-induced hemodynamic changes were referenced to a consistent baseline level, the mean value of the 30-second baseline was subtracted from the entire time series.

For the affective state, hemodynamic responses were averaged across the Positive and Negative conditions for each participant. Mean changes in hemoglobin concentration were calculated for the during- and post-stimulation phases, with 5% Winsorization applied to attenuate outlier effects. Moreover, state-specific time courses were constructed by averaging HbO and HbR changes over consecutive 20-second intervals from audio onset through the end of the post-stimulation phase, yielding a total of 20 time windows (w_1_-w_20_) referenced to baseline. These values were subsequently used in the statistical analyses.

### 2.4 Statistical analysis

Demographic characteristics were summarized using descriptive statistics. Subjective ratings (SAM and VAS pain scores) were compared using one-way repeated-measures analysis of variance (RM ANOVA), followed by Bonferroni-corrected post-hoc pairwise comparisons where appropriate.

For the primary analysis, two-way RM ANOVAs were performed separately on ΔHbO and ΔHbR, with State (control vs. affective) and Time Phase (during-stimulation vs. post-stimulation) as within-subject factors, to investigate the overall impact of auditory-induced affective on iTBS-evoked hemodynamic responses. To control for the familywise error rate across two hemoglobin metrics, the significance threshold was set at p < 0.025 (i.e., 0.05/2). When a significant interaction was observed, post-hoc tests with Bonferroni correction were conducted to evaluate simple main effects. The above statistical analyses were performed using SPSS (version 27.0.1; IBM Corp., Armonk, NY, USA).

To characterize state-specific temporal trajectories, linear mixed-effects models (LMM) were implemented in R (version 4.6.0; The R Foundation for Statistical Computing, Vienna, Austria) using the lmerTest package. Fixed effects included State (control, affective), Time Window (baseline and w_1_-w_20_), and their interaction, alongside random intercepts for participants. Model-estimated marginal means were evaluated to compare each time window against baseline within each state. False discovery rate (FDR) correction was applied across the comparisons, with q < 0.05 considered statistically significant.

## 3. Results

### 3.1 Demographics and subjective ratings

All thirty-six enrolled participants completed the study without any dropouts, and all collected data met the predetermined quality requirements. Participant characteristics are summarized in Table 1. The auditory stimuli successfully manipulated participants’ affective states. For the SAM valence ratings, one-way RM ANOVA revealed a significant difference across the three auditory conditions (F_2,70_ = 55.23, p < 0.001, partial η² = 0.612). Bonferroni-corrected post-hoc pairwise comparisons confirmed that all three conditions differed significantly from each other (p_corr_ < 0.001), with the Negative condition inducing unpleasant valence (3.75 ± 1.50), the Positive condition inducing pleasant valence (7.06 ± 1.26), and the Neutral condition remaining near the scale midpoint (5.39 ± 1.50).

**Table 1.** Demographic information of the study participants.

| Characteristics | Participants (N = 36, Sessions = 108) |
|---|---|
| Age, years, mean (SD) | 23.94 (4.47) |
| Gender, N of female (%) | 18 (50%) |
| Education years, mean (SD) | 15.50 (1.83) |
| Ethnicity, N of Han Chinese (%) | 36 (100%) |
| PHQ-9, mean (SD) | 1.86 (1.44) |
| Head circumference, mean (SD) | 56.27 (1.59) |
| RMT with cap, mean (SD) | 68.67 (11.58) |

Regarding physical tolerability, iTBS was significantly better tolerated during the Positive condition compared to the other states. One-way RM ANOVA confirmed a significant main effect of condition on VAS pain scores (F_2,70_ = 4.94, p = 0.010, partial η² = 0.124), with post-hoc tests demonstrating reduced pain in the Positive condition (3.24 ± 1.79) relative to both the Neutral (3.86 ± 1.98, p_corr_ =0.011) and Negative (3.92 ± 1.90, p_corr_ = 0.027) conditions. One participant reported mild anxious feelings after the Negative condition, which resolved spontaneously. No other adverse events occurred. A summary of all subjective ratings is provided in Table 2.

**Table 2.** Summary of self-reported SAM and VAS pain ratings.

|  | Neutral | Positive | Negative | F<br>(df1, df2) | P value | partial<br>η<sup>2</sup> | Post-hoc comparisons |
|---|---|---|---|---|---|---|---|
| Valence (SAM), pleasure, mean (SD) | 5.39<br>(1.50) | 7.06<br>(1.26) | 3.75<br>(1.50) | 55.23<br>(2,70) | **< 0.001** | 0.612 | Negative < Neutral < Positive,<br>**all p<sub>corr</sub> < 0.001** |
| Arousal (SAM), excited, mean (SD) | 4.78<br>(1.74) | 4.67<br>(1.84) | 5.53<br>(1.77) | 3.10<br>(2,70) | 0.051 | 0.081 | / |
| Dominance (SAM), in control, mean (SD) | 5.86<br>(1.82) | 7.25<br>(1.34) | 4.86<br>(1.89) | 21.21<br>(2,70) | **< 0.001** | 0.377 | Neutral < Positive, **p<sub>corr</sub> < 0.001**;<br>Negative < Positive, **p<sub>corr</sub> < 0.001**;<br>Negative < Neutral, **p<sub>corr</sub> = 0.033;** |
| Pain during iTBS (VAS), mean (SD) | 3.86<br>(1.98) | 3.24<br>(1.79) | 3.92<br>(1.90) | 4.94<br>(2,70) | **0.010** | 0.124 | Positive < Neutral, **p<sub>corr</sub> = 0.011**;<br>Positive < Negative, **p<sub>corr</sub> = 0.027**; |

### 3.2 Hemodynamic responses

#### Effect of Affective States

The two-way RM ANOVA results are shown in Table 3 and visualized in Figure 2. For ΔHbO, no significant State × Time Phase interaction (p = 0.087) and no significant main effect of Time Phase (p = 0.957) were observed. However, a significant main effect of State was observed (F_1,35_ = 6.52, p = 0.015, partial η² = 0.157), which survived Bonferroni adjustment. Exploratory paired-samples t-tests indicated that ΔHbO was significantly higher in the control state than in the affective state both during the stimulation phase (mean difference = 0.15, 95% CI [0.03, 0.27], t_(35)_ = 2.48, p = 0.018, Cohen’s d = 0.41) and in the post-stimulation phase (mean difference = 0.10, 95% CI [0.02, 0.18], t_(35)_ = 2.55, p = 0.015, Cohen’s d = 0.43). To test whether this observed difference was target-site specific rather than a generalized affect-driven or systemic physiological artifact, we compared HbO responses at the stimulated (left) and contralateral (right) DLPFC in the Supplementary Material (Section A).

**Figure 2.**
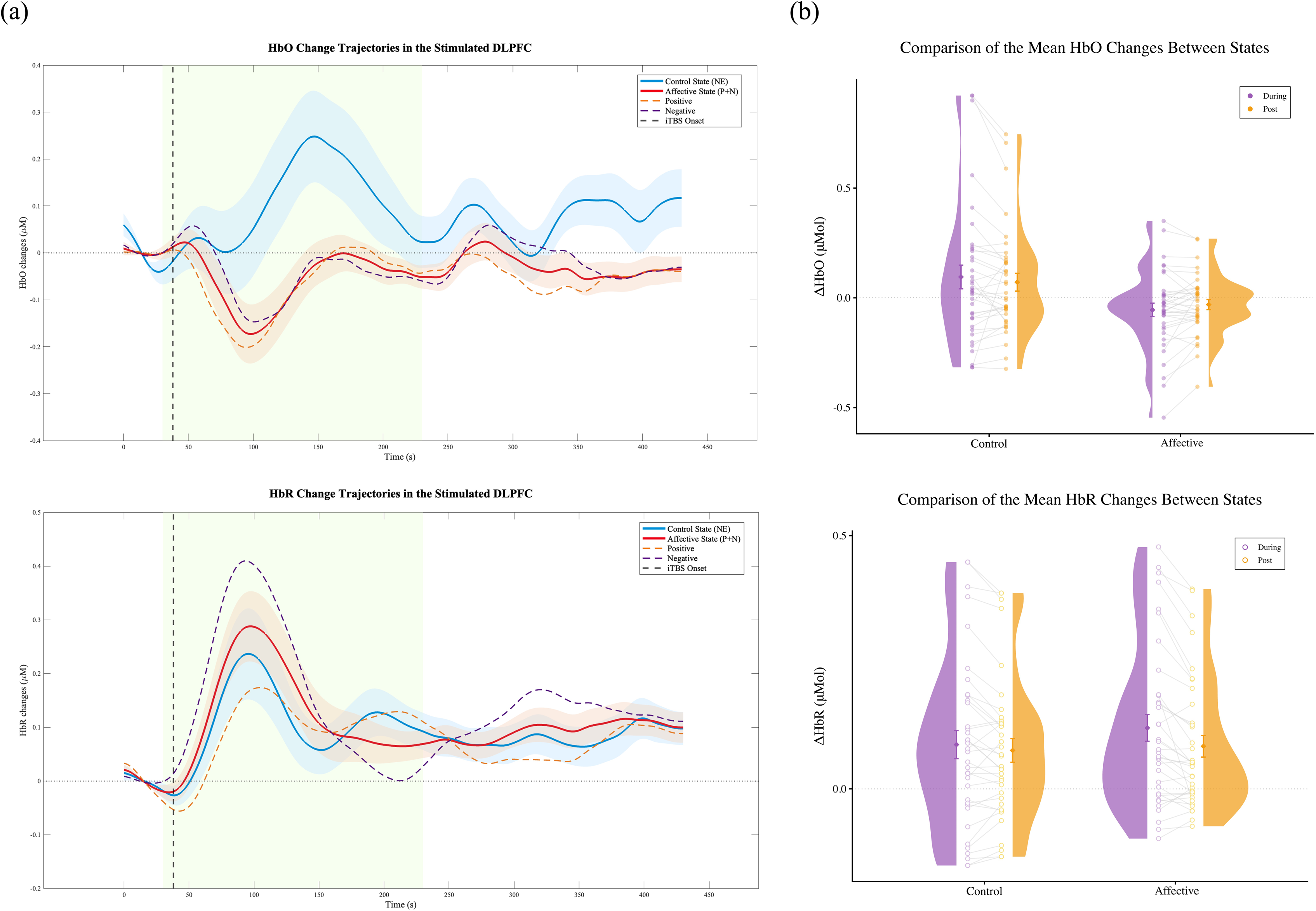
Hemodynamic responses in the left DLPFC. **(a)** Real-time trajectories of HbO and HbR changes across two states. Solid lines and corresponding shadings denote the mean ± 1 standard error of the mean (SEM); colored dashed lines show the condition means, representing the positive and negative conditions that comprise the affective state. The green background highlights the during-stimulation phase (200 s), with the vertical dashed line marking iTBS onset. **(b)** Mean ΔHbO and ΔHbR comparisons between control and affective states during and post-stimulation. The during-stimulation phase is shown in purple, and the post-stimulation phase in orange. Light gray lines connect individual participant data points (n = 36) across the two phases. Half-violin plots display participant distributions, with diamonds and error bars indicating the mean ± SEM.

**Table 3.** Two-way RM ANOVA results for hemoglobin changes in the stimulated DLPFC across states and time phases.

| Hb Type | State |  |  | Time Phase |  |  | State × Time Phase Interaction |  |  |
|---|---|---|---|---|---|---|---|---|---|
|  | F (df□, df□) | p<sup>*</sup> | partial η² | F (df□, df□) | p<sup>*</sup> | partial η² | F (df□, df□) | p<sup>*</sup> | partial η² |
| HbO | 6.518 (1,35) | **0.015** | 0.157 | 0.003 (1,35) | 0.957 | < 0.001 | 3.104 (1,35) | 0.087 | 0.081 |
| HbR | 0.653 (1, 35) | 0.425 | 0.018 | 14.378 (1,35) | **0.001** | 0.291 | 5.996 (1, 35) | **0.019** | 0.146 |

<sup>\*</sup> Significant adjusted to p < 0.025 (i.e., 0.05/2, based on Bonferroni adjustments)

For ΔHbR, the results revealed a significant State × Time Phase interaction effect (F_1,35_ = 6.00, p = 0.019, partial η² = 0.146) and a main effect of Time Phase (F_1,35_ = 14.38, p = 0.001, partial η² = 0.291), whereas the main effect of State was not significant (p = 0.425). Decomposing the significant interaction via simple main effects analyses showed that in the affective state, ΔHbR during the stimulation phase was significantly higher than in the post-stimulation phase (mean difference = 0.04, 95% CI [0.02, 0.05], t(35) = 4.04, p < 0.001, Cohen’s d = 0.67). In the control state, ΔHbR did not differ significantly between the two phases (p = 0.111). No significant differences in ΔHbR were observed between states in either time phase (all p > 0.05).

Additional exploration of the temporal and morphological characteristics (amplitude, latency, and duration) of the hemodynamic responses between the Positive and Negative conditions is provided in the Supplementary Material (Section B).

#### Time-course Analysis

Figure 3 presents the hemodynamic time courses for both the control and affective states across baseline and the successive 20-second windows (w_1_-w_20_). The LMM results indicated no significant HbO deviations from baseline across any individual time window in either state (all q > 0.05). Regarding HbR, both states followed a similar trajectory – rising initially and then declining. Nevertheless, the control state exhibited its first significant change at w_3_ (mean difference = 0.18, 95% CI [0.10, 0.27], q = 0.015, Cohen’s d = 0.93) and peaked at w_4_ (mean difference = 0.23, 95% CI [0.14, 0.31], q < 0.001, Cohen’s d = 1.15), with the effect dissipating quickly thereafter. By contrast, the affective state demonstrated a stronger and more sustained HbR elevation in response to iTBS, spanning w_3_ (mean difference = 0.23, 95% CI [0.15, 0.31], q < 0.001, Cohen’s d = 1.17), w_4_ (mean difference = 0.28, 95% CI [0.20, 0.37], q < 0.001, Cohen’s d = 1.43) and w_5_ (mean difference = 0.23, 95% CI [0.15, 0.32], q < 0.001, Cohen’s d = 1.18) relative to baseline. Full pairwise comparisons against baseline across all time windows are summarized in Table 4.

**Figure 3.**
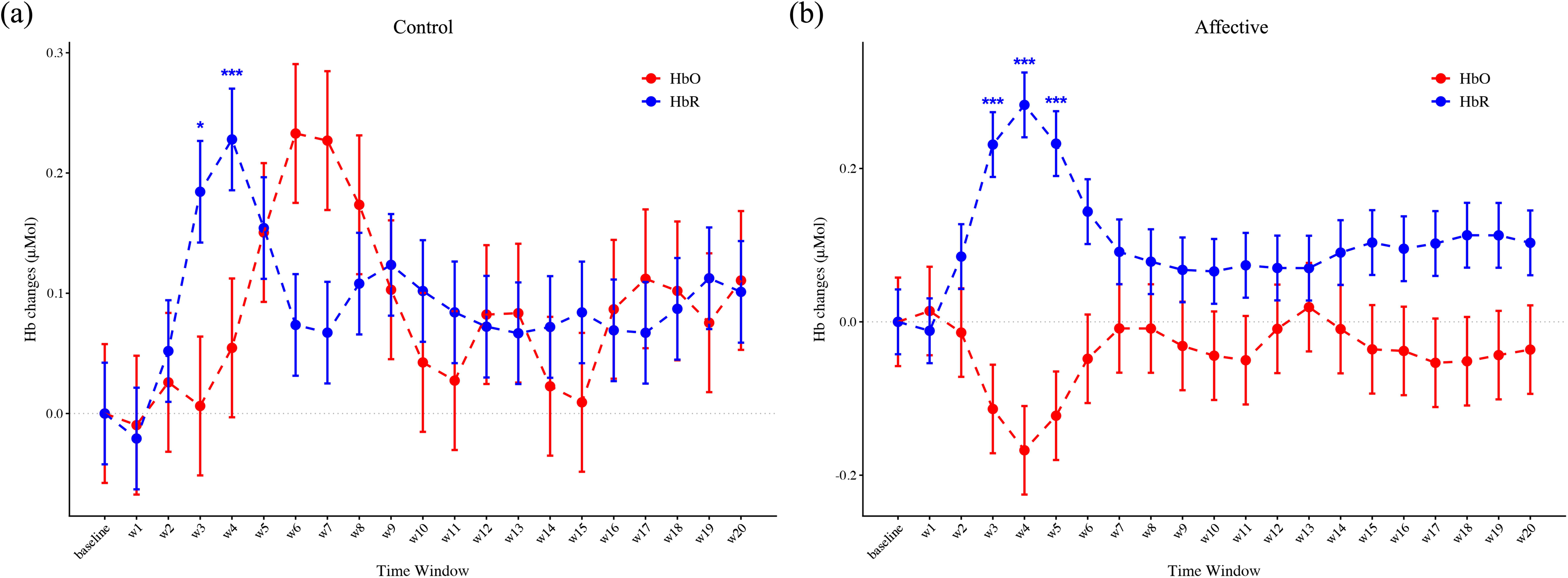
Time course analysis of hemodynamic change profiles. Hb changes are plotted across baseline and 20 subsequent time windows (w_1_ to w_20_); each window represents a 20-second average from audio onset. Panels display results for **(a)** the control state and **(b)** the affective state. Points represent estimated marginal means from the LMM, with error bars indicating the standard error. Significant deviations from baseline following FDR correction within each state and chromophore are indicated by asterisks (*q < 0.05, ***q < 0.0001).

**Table 4.** LMM analysis of hemoglobin changes in the stimulated DLPFC across time windows relative to baseline.

| Time Window | HbO |  |  |  |  |  | HbR |  |  |  |  |  |
|---|---|---|---|---|---|---|---|---|---|---|---|---|
|  | Control State |  |  | Affective State |  |  | Control State |  |  | Affective State |  |  |
|  | Mean Difference | q | Cohen’s d | Mean Difference | q | Cohen’s d | Mean Difference | q | Cohen’s d | Mean Difference | q | Cohen’s d |
| w1 | -0.01 | 1.000 | -0.03 | 0.01 | 1.000 | 0.05 | -0.02 | 0.999 | -0.11 | -0.01 | 1.000 | -0.06 |
| w2 | 0.03 | 1.000 | 0.09 | -0.01 | 1.000 | -0.05 | 0.05 | 0.970 | 0.26 | 0.09 | 0.814 | 0.43 |
| w3 | 0.01 | 1.000 | 0.02 | -0.11 | 1.000 | -0.38 | 0.18 | **0.015** | 0.93 | 0.23 | **< 0.001** | 1.17 |
| w4 | 0.05 | 1.000 | 0.18 | -0.17 | 1.000 | -0.55 | 0.23 | **< 0.001** | 1.15 | 0.28 | **< 0.001** | 1.43 |
| w5 | 0.15 | 1.000 | 0.50 | -0.12 | 1.000 | -0.41 | 0.15 | 0.106 | 0.78 | 0.23 | **< 0.001** | 1.18 |
| w6 | 0.23 | 0.238 | 0.77 | -0.05 | 1.000 | -0.16 | 0.07 | 0.869 | 0.37 | 0.14 | 0.164 | 0.73 |
| w7 | 0.23 | 0.238 | 0.75 | -0.01 | 1.000 | -0.03 | 0.07 | 0.869 | 0.34 | 0.09 | 0.757 | 0.46 |
| w8 | 0.17 | 1.000 | 0.57 | -0.01 | 1.000 | -0.03 | 0.11 | 0.774 | 0.55 | 0.08 | 0.835 | 0.40 |
| w9 | 0.10 | 1.000 | 0.34 | -0.03 | 1.000 | -0.10 | 0.12 | 0.544 | 0.63 | 0.07 | 0.835 | 0.34 |
| w10 | 0.04 | 1.000 | 0.14 | -0.04 | 1.000 | -0.15 | 0.10 | 0.774 | 0.52 | 0.07 | 0.835 | 0.33 |
| w11 | 0.03 | 1.000 | 0.09 | -0.05 | 1.000 | -0.17 | 0.08 | 0.869 | 0.43 | 0.07 | 0.835 | 0.37 |
| w12 | 0.08 | 1.000 | 0.27 | -0.01 | 1.000 | -0.03 | 0.07 | 0.869 | 0.37 | 0.07 | 0.835 | 0.36 |
| w13 | 0.08 | 1.000 | 0.28 | 0.02 | 1.000 | 0.06 | 0.07 | 0.869 | 0.34 | 0.07 | 0.835 | 0.35 |
| w14 | 0.02 | 1.000 | 0.08 | -0.01 | 1.000 | -0.03 | 0.07 | 0.869 | 0.36 | 0.09 | 0.757 | 0.46 |
| w15 | 0.01 | 1.000 | 0.03 | -0.04 | 1.000 | -0.12 | 0.08 | 0.869 | 0.43 | 0.10 | 0.661 | 0.52 |
| w16 | 0.09 | 1.000 | 0.29 | -0.04 | 1.000 | -0.13 | 0.07 | 0.869 | 0.35 | 0.10 | 0.757 | 0.48 |
| w17 | 0.11 | 1.000 | 0.37 | -0.05 | 1.000 | -0.18 | 0.07 | 0.869 | 0.34 | 0.10 | 0.661 | 0.52 |
| w18 | 0.10 | 1.000 | 0.34 | -0.05 | 1.000 | -0.17 | 0.09 | 0.869 | 0.44 | 0.11 | 0.625 | 0.57 |
| w19 | 0.08 | 1.000 | 0.25 | -0.04 | 1.000 | -0.14 | 0.11 | 0.762 | 0.57 | 0.11 | 0.625 | 0.57 |
| w20 | 0.11 | 1.000 | 0.37 | -0.04 | 1.000 | -0.12 | 0.10 | 0.774 | 0.51 | 0.10 | 0.661 | 0.52 |

## 4. Discussion

This study indicates that affective auditory stimulation interleaved with iTBS differentially modulated both fNIRS measures. Whereas HbO levels were significantly higher in the control state than in the affective state across both phases, HbR displayed distinct temporal trajectories across states, despite exhibiting a shared initial post-onset increase followed by a return toward baseline.

The contrasting HbO patterns and significant differences observed across both phases between the two states indicate that the neuromodulatory effects of iTBS are influenced by audio-induced affect. Notably, the decrease in ΔHbO during stimulation in the affective state parallels findings from previous concurrent fNIRS studies employing higher-intensity (90% RMT) iTBS at rest in both healthy individuals and patients with MDD^28,29,40,49^. A commonly proposed explanation for this ΔHbO reduction under excitatory stimulation is that high-frequency stimulation activates the sympathetic nervous system, prompting vasoconstriction in cerebral resistance vessels that overrides blood pressure-induced increases in CBF^50^. Aligning with these reports, our findings suggest that affective processing can modulate the hemodynamic response, eliciting localized HbO changes that resemble patterns typically observed at higher stimulation intensities.

Beyond HbO, we supplemented our interpretation with HbR as a complementary marker of systemic physiological changes associated with iTBS^51^, which also reflects the heightened metabolic demands underlying neuronal activity and synaptic transmission^52^. Integrating both chromophores, the control state exhibited a concurrent rise in both ΔHbO and ΔHbR at the target site during stimulation. Although such a pattern diverging from classic NVC has not been reported in other TMS studies, it is consistent with findings from a 50 Hz cortical electrical stimulation study^53^. The authors reported the same hemodynamic pattern and attributed it to the “fast-oxygen response in capillary event”^54^, a phenomenon wherein high-frequency stimulation drives intense neural activity that accelerates capillary oxygen exchange^53^. Hence, despite a marked increase in local blood volume and an augmented HbO supply, transient HbR accumulation outpaces washout, resulting in this co-elevation pattern. Also, cellular-level work using focused transcranial ultrasound stimulation (TUS) – an excitatory NIBS modality – demonstrated a concurrent increase in HbR that surpassed that of HbO during stimulation in murine brains^55^. This suggests that localized activation may trigger excessive local oxygen consumption, or that non-selective stimulation parameters exert a stronger effect on inhibitory neuronal populations than on excitatory ones, ultimately shaping the hemodynamic pattern^55^. In contrast, iTBS administered under the affective state induced a decrease in HbO alongside an increase in HbR, reversing the typical hemodynamic response associated with the left DLPFC activation during affective processing^56^. Drawing on the “vascular stealing” hypothesis^57^, this inverted hemodynamic pattern observed at the target site may reflect competition for CBF between concurrent affective processing networks and iTBS-induced NVC response. The higher metabolic demand of widely distributed affective networks could actively divert blood supply away from the left DLPFC to other related regions.

Furthermore, fine-grained temporal trajectories characterized the trends of each state. Relative to baseline, HbO showed no statistically significant changes at any time window in either state. For HbR, the rapid rise and dissipation observed at w_2_ and w_3_ in the control state reflected an initial transient response to stimulation and then adaptation. When paired with affective processing, the HbR trajectory exhibited a smoother, more lasting profile, maintaining significant baseline elevations across three consecutive time windows. This finding converges with our primary outcome, in which the two time phases differed significantly only within the affective state. Similar hemodynamic features have been found in patients with MDD, who experience a persistent negative affective state and display heightened HbR responses during iTBS compared to healthy controls^40,49^. Notably, participants reported significantly less pain during iTBS when paired with positive valence induction, offering valuable insight into strategies for reducing treatment-related discomfort in clinical applications.

### 4.1 Limitations and implications for future direction

Several limitations warrant consideration for future affective state-dependent neuromodulation research. First, systemic physiological parameters were not monitored in the present study to prioritize participant comfort. However, concurrent blood pressure recording is recommended to better dissociate non-neural contributions^58^, while heart rate variability (HRV) – a validated objective biomarker of autonomic affect regulation^59,60^ – could complement subjective ratings and index real-time affective fluctuations. Second, other affective dimensions and acoustic properties of the induction audio should be more strictly controlled to minimize confounding influences^15^. Relatedly, our experimental design induced affect solely along a dimensional valence framework, which may oversimplify multifaceted affective experiences^61^. Future work should incorporate distinct affective categories. Additionally, because audio clip selection was constrained by valence ratings, the number of clips and repetition times differed between the affective and control states, representing a methodological discrepancy. Third, this study was conducted in healthy adults, whereas clinical populations often present with persistent affective disturbances (e.g., chronic anhedonia and affective blunting in patients with MDD), exhibiting a negative baseline alongside deficits in affect regulation^62,63^. Determining whether pre-conditioning affective state or delivering concurrent affective modulation yields superior therapeutic efficacy will be critical for developing affective-dependent strategies. Finally, elucidating the underlying NVC mechanisms across macroscopic and microscopic scales will be essential to link these hemodynamic responses to specific stimulation parameters and clinical outcomes.

## 5. Conclusion

Our findings provide the first evidence that auditory-induced affective state modulates left DLPFC engagement in response to iTBS. Overall state differences were reflected in HbO, with the concentration changes being lower in the affective state in comparison with the control state. Although HbR showed an initial increase followed by a decline in both states, fine-grained temporal dynamics captured the greater and more sustained changes when stimulation was combined with affective processing elicited by audio stimuli. Additionally, pairing iTBS with positive valence effectively mitigated stimulation-related discomfort. Together, these findings highlight the potential value of accounting for affective state in neuromodulation strategies; replicating and extending them to clinical cohorts will be essential for translating affective modulation into enhanced treatment tolerability and efficacy.

## Authorship contribution statement

**Nancy M.X.Y. Shi:** Conceptualization, Methodology, Formal analysis, Data curation, Investigation, Writing - Original Draft, Writing – Review & Editing, Visualization. **Alvin H.P. Tang:** Conceptualization, Methodology, Data curation, Investigation, Writing – Review & Editing. **Minxia Jin:** Methodology, Writing – Review & Editing. **Adam W.L. Xia:** Methodology, Writing – Review & Editing. **Rebecca L.D. Kan:** Methodology, Writing – Review & Editing. **Bella B.B. Zhang:** Methodology, Writing – Review & Editing. **Penny P. Qin:** Writing – Review & Editing. **Tim T.Z. Lin:** Methodology. **Sharie X. Wang:** Investigation. **Vera W.M. Lam:** Investigation. **Wanda M.W. Chau:** Investigation. **Frank Padberg:** Writing – Review & Editing. **Juha Silvanto:** Writing – Review & Editing. **Georg S. Kranz:** Conceptualization, Methodology, Data curation, Writing – Review & Editing, Resources, Supervision, Funding acquisition.

## Data availability

The data are available from the corresponding author upon reasonable request.

## Acknowledgement

The authors would like to acknowledge the support from the University Research Facility in Behavioral and Systems Neuroscience (UBSN). We also thank Chuanliang Han for his valuable comments on this study.

## Declarations of generative AI and AI-assisted technologies in the manuscript preparation process

During the preparation of this work, the author(s) used Gemini 3.8 Flash to polish the wording. After using this tool/service, the author(s) reviewed and edited the content as needed and take(s) full responsibility for the content of the published article.

## Reference

1. Desarkar P, Vicario C, Soltanlou M. Non-invasive brain stimulation in research and therapy. Editorial Material. Sci Rep. Nov 26 2024;14(1):4. 29334. doi:10.1038/s41598-024-79039-1

2. Klomjai W, Katz R, Lackmy-Vallée A. Basic principles of transcranial magnetic stimulation (TMS) and repetitive TMS (rTMS). Ann Phys Rehabil Med. Sep 2015;58(4):208–213. doi:10.1016/j.rehab.2015.05.005

3. Kishi T, Sakuma K, Matsuda Y, Kito S, Iwata N. Intermittent theta burst stimulation vs. high-frequency repetitive transcranial magnetic stimulation for major depressive disorder: A systematic review and meta-analysis. Review. Psychiatry Res. Oct 2023;328:3. 115452. doi:10.1016/j.psychres.2023.115452

4. Lan X, Yang X, Qin Z, et al. Efficacy and safety of intermittent theta burst stimulation versus high-frequency repetitive transcranial magnetic stimulation for patients with treatment-resistant depression: a systematic review. Review. Front Psychiatry. Jul 2023;14:8. 1244289. doi:10.3389/fpsyt.2023.1244289

5. Blumberger D, Vila-Rodriguez F, Thorpe K, et al. Effectiveness of theta burst versus high-frequency repetitive transcranial magnetic stimulation in patients with depression (THREE-D): a randomised non-inferiority trial. Lancet. Apr 2018;391(10131):1683–1692. doi:10.1016/s0140-6736(18)30295-2

6. Friedman N, Robbins T. The role of prefrontal cortex in cognitive control and executive function. Review. Neuropsychopharmacology. Jan 2022;47(1):72–89. doi:10.1038/s41386-021-01132-0

7. Zhang G, Yao L, Zhang H, Long Z, Zhao X. Improved Working Memory Performance through Self-Regulation of Dorsal Lateral Prefrontal Cortex Activation Using Real-Time fMRI. PLoS One. Aug 2013;8(8):9.e73735. doi:10.1371/journal.pone.0073735

8. Hwang J, Kim S, Park C, Bang S, Kim S. Acute high-frequency rTMS of the left dorsolateral prefrontal cortex and attentional control in healthy young men. Brain Res. May 2010;1329:152–158. doi:10.1016/j.brainres.2010.03.013

9. White L, Makhoul W, Teferi M, Sheline Y, Balderston N. The role of dlPFC laterality in the expression and regulation of anxiety. Neuropharmacology. Feb 2023;224:11. 109355. doi:10.1016/j.neuropharm.2022.109355

10. Kishi T, Ikuta T, Sakuma K, et al. Theta burst stimulation for depression: a systematic review and network and pairwise meta-analysis. Review. Mol Psychiatr. Dec 2024;29(12):3893–3899. doi:10.1038/s41380-024-02630-5

11. Ning MH, Sun H, Passera B, et al. Complexity of resting cortical activity predicts neurophysiological responses to theta-burst stimulation but fails to generalize: A rigorous machine-learning approach. PLoS Comput Biol. Apr 2026;22(4):e1014154. doi:10.1371/journal.pcbi.1014154

12. Ozdemir RA, Boucher P, Fried PJ, et al. Reproducibility of cortical response modulation induced by intermittent and continuous theta-burst stimulation of the human motor cortex. Brain Stimul. Jul–Aug 2021;14(4):949–964. doi:10.1016/j.brs.2021.05.013

13. Silvanto J, Pascual-Leone A. State-dependency of transcranial magnetic stimulation. Review. Brain Topogr. Sep 2008;21(1):1–10. doi:10.1007/s10548-008-0067-0

14. Sack AT, Paneva J, Küthe T, et al. Target Engagement and Brain State Dependence of Transcranial Magnetic Stimulation: Implications for Clinical Practice. Biological Psychiatry. 2024/03/15/2024;95(6):536–544. 10.1016/j.biopsych.2023.09.011

15. Schutter DJLG, Smits F, Klaus J. Mind matters: A narrative review on affective state-dependency in non-invasive brain stimulation. International Journal of Clinical and Health Psychology. 2023/07/01/2023;23(3):100378. 10.1016/j.ijchp.2023.100378

16. Wu T, Yu Q, Zhu X, et al. Embracing Internal States: A Review of Optimization of Repetitive Transcranial Magnetic Stimulation for Treating Depression. Neurosci Bull. May 2025;41(5):866–880. doi:10.1007/s12264-024-01347-3

17. Lindquist K, Satpute A, Wager T, Weber J, Barrett L. The Brain Basis of Positive and Negative Affect: Evidence from a Meta-Analysis of the Human Neuroimaging Literature. Cereb Cortex. May 2016;26(5):1910–1922. doi:10.1093/cercor/bhv001

18. Viinikainen M, Kätsyri J, Sams M. Representation of perceived sound valence in the human brain. Human Brain Mapping. 2012;33(10):2295–2305. 10.1002/hbm.21362

19. Hou Y, Cao X, Zhang Y, Shi H. fNIRS evidence for valence-dependent prefrontal activation moderating resilience to depression. J Affect Disord. Feb 2026;395:9. 120771. doi:10.1016/j.jad.2025.120771

20. Nejati V, Majdi R, Salehinejad M, Nitsche M. The role of dorsolateral and ventromedial prefrontal cortex in the processing of emotional dimensions. Sci Rep. Jan 2021;11(1):12. 1971. doi:10.1038/s41598-021-81454-7

21. Lee Y, Seo Y, Lee Y, Lee D. Dimensional emotions are represented by distinct topographical brain networks. International Journal of Clinical and Health Psychology. Oct–Dec 2023;23(4):12. 100408. doi:10.1016/j.ijchp.2023.100408

22. Pinti P, Tachtsidis I, Hamilton A, et al. The present and future use of functional near-infrared spectroscopy (fNIRS) for cognitive neuroscience. Review. Ann NY Acad Sci. Mar 2020;1464(1):5–29. doi:10.1111/nyas.13948

23. Parks NA. Concurrent application of TMS and near-infrared optical imaging: methodological considerations and potential artifacts. Review. Front Hum Neurosci. 2013–September–19 2013;Volume 7 -2013doi:10.3389/fnhum.2013.00592

24. Curtin A, Tong S, Sun J, Wang J, Onaral B, Ayaz H. A Systematic Review of Integrated Functional Near-Infrared Spectroscopy (fNIRS) and Transcranial Magnetic Stimulation (TMS) Studies. Review. Front Neurosci. Feb 2019;13:23. 84. doi:10.3389/fnins.2019.00084

25. Scarapicchia V, Brown C, Mayo C, Gawryluk J. Functional Magnetic Resonance Imaging and Functional Near-Infrared Spectroscopy: Insights from Combined Recording Studies. Review. Front Hum Neurosci. Aug 2017;11:12. 419. doi:10.3389/fnhum.2017.00419

26. Rahman MA, Siddik AB, Ghosh TK, Khanam F, Ahmad M. A Narrative Review on Clinical Applications of fNIRS. Journal of Digital Imaging. 2020/10/01 2020;33(5):1167–1184. doi:10.1007/s10278-020-00387-1

27. Yu Q, Yuan Q, Song Y, et al. Functional near-infrared spectroscopy (fNIRS) in medical research: applications, opportunities, and challenges. Chinese Medicine. 2026/06/18 2026;21(1):169. doi:10.1186/s13020-026-01409-w

28. Kan RL, Zhang BB, Lin TT, et al. Sex differences in brain excitability revealed by concurrent iTBS/fNIRS. Asian journal of psychiatry. 2024;96:104043.

29. Xia A, Jin M, Zhang B, et al. Investigating the hemodynamic response to iTBS of the left DLPFC: A concurrent iTBS/fNIRS study. Brain Stimul. Mar–Apr 2025;18(2):235–245. doi:10.1016/j.brs.2025.02.008

30. Rossi S, Hallett M, Rossini P, Pascual-Leone A. Screening questionnaire before TMS: An update. Letter. Clin Neurophysiol. Aug 2011;122(8):1686–1686. doi:10.1016/j.clinph.2010.12.037

31. Yang N, Waddington G, Adams R, Han J. Translation, cultural adaption, and test-retest reliability of Chinese versions of the Edinburgh Handedness Inventory and Waterloo Footedness Questionnaire. Laterality. 2018 2018;23(3):255–273. doi:10.1080/1357650x.2017.1357728

32. Wang W, Bian Q, Zhao Y, et al. Reliability and validity of the Chinese version of the Patient Health Questionnaire (PHQ-9) in the general population. Gen Hosp Psych. Sep 2014;36(5):539–544. doi:10.1016/j.genhosppsych.2014.05.021

33. Kudo D, Nito M, Kikuchi A, Tanabe S, Yamaguchi T. Individualized Beta Frequency Intermittent Theta Burst Stimulation Enhances Corticospinal Plasticity in Healthy Adults. Neuromodulation: Technology at the Neural Interface. 2026/02/01/2026;29(2):298–305. 10.1016/j.neurom.2025.10.069

34. Kan RL, Lin TT, Zhang BB, et al. Moderators of stimulation-induced neural excitability in the left DLPFC: A concurrent iTBS/fNIRS case study. Brain Stimulation: Basic, Translational, and Clinical Research in Neuromodulation. 2023;16(5):1445–1447.

35. Yang W, Makita K, Nakao T, et al. Affective auditory stimulus database: An expanded version of the International Affective Digitized Sounds (IADS-E). Behav Res Methods. Aug 2018;50(4):1415–1429. doi:10.3758/s13428-018-1027-6

36. Bradley MM, Lang PJ. Measuring emotion: the self-assessment manikin and the semantic differential. Journal of behavior therapy and experimental psychiatry. 1994;25(1):49–59.

37. Huang Y, Edwards M, Rounis E, Bhatia K, Rothwell J. Theta burst stimulation of the human motor cortex. Neuron. Jan 20 2005;45(2):201–206. doi:10.1016/j.neuron.2004.12.033

38. Nettekoven C, Volz LJ, Kutscha M, et al. Dose-dependent effects of theta burst rTMS on cortical excitability and resting-state connectivity of the human motor system. J Neurosci. May 14 2014;34(20):6849–59. doi:10.1523/jneurosci.4993-13.2014

39. Zhang BBB, Kan RLD, Giron CG, Lin TTZ, Yau SY, Kranz GS. Dose-response relationship between iTBS and prefrontal activation during executive functioning: A fNIRS study. Front Psychiatry. 2022;13:1049130. doi:10.3389/fpsyt.2022.1049130

40. Jin M, Xia A, Chau W, et al. Acute prefrontal hemodynamic responses to intermittent theta burst stimulation correlate with current depression and episode recurrence: A cross-sectional study. ; Early Access. Psychiatry Clin Neurosci. 2026:10. doi:10.1111/pcn.70066

41. Huppert T, Diamond S, Franceschini M, Boas D. HomER: a review of time-series analysis methods for near-infrared spectroscopy of the brain. ; Proceedings Paper. Appl Optics. Apr 1 2009;48(10):D280–D298. doi:10.1364/ao.48.00d280

42. Yücel M, Lühmann A, Scholkmann F, et al. Best practices for fNIRS publications. Neurophotonics. Jan–Mar 2021;8(1):34. 012101. doi:10.1117/1.NPh.8.1.012101

43. Kocsis L, Herman P, Eke A. The modified Beer-Lambert law revisited. Phys Med Biol. Mar 7 2006;51(5):N91–N98. doi:10.1088/0031-9155/51/5/n02

44. Yücel M, Selb J, Aasted C, et al. Short separation regression improves statistical significance and better localizes the hemodynamic response obtained by near-infrared spectroscopy for tasks with differing autonomic responses. Neurophotonics. Jul–Sep 2015;2(3):10. 035005. doi:10.1117/1.NPh.2.3.035005

45. Zhang H, Zhang Y, Lu C, Ma S, Zang Y, Zhu C. Functional connectivity as revealed by independent component analysis of resting-state fNIRS measurements. Neuroimage. Jul 1 2010;51(3):1150–1161. doi:10.1016/j.neuroimage.2010.02.080

46. Zhu H, Xu J, Li J, et al. Decreased functional connectivity and disrupted neural network in the prefrontal cortex of affective disorders: A resting-state fNIRS study. J Affect Disord. Oct 15 2017;221:132–144. doi:10.1016/j.jad.2017.06.024

47. Molavi B, Dumont G. Wavelet-based motion artifact removal for functional near-infrared spectroscopy. Physiol Meas. Feb 2012;33(2):259–270. doi:10.1088/0967-3334/33/2/259

48. Pinti P, Scholkmann F, Hamilton A, Burgess P, Tachtsidis I. Current Status and Issues Regarding Pre-processing of fNIRS Neuroimaging Data: An Investigation of Diverse Signal Filtering Methods Within a General Linear Model Framework. Front Hum Neurosci. Jan 11 2019;12:21. 505. doi:10.3389/fnhum.2018.00505

49. Kan RLD, Tang AHP, Jin M, et al. Cross-sectional and longitudinal analysis of prefrontal intermittent theta-burst stimulation-evoked hemodynamic responses in major depressive disorder. Translational Psychiatry. 2026/07/17 2026;doi:10.1038/s41398-026-04271-4

50. Vernieri F, Maggio P, Tibuzzi F, et al. High frequency repetitive transcranial magnetic stimulation decreases cerebral vasomotor reactivity. Clin Neurophysiol. 2009/06/01/2009;120(6):1188–1194. 10.1016/j.clinph.2009.03.021

51. Tachtsidis I, Scholkmann F. False positives and false negatives in functional near-infrared spectroscopy: issues, challenges, and the way forward. Neurophotonics. 2016 Jul (Epub 2016 Mar 2016;3(3):030401. doi:10.1117/1.NPh.3.3.030401

52. Zhao YN, Zhang XY, Huang YY, Wu JC, Bi X. Intermittent theta burst stimulation enhances prefrontal activation and connectivity: evidence from fNIRS. Front Hum Neurosci. 2025;19:1608502. doi:10.3389/fnhum.2025.1608502

53. Sato Y, Oishi M, Fukuda M, Fujii Y. Hemodynamic and electrophysiological connectivity in the language system: Simultaneous near-infrared spectroscopy and electrocorticography recordings during cortical stimulation. Brain and Language. 2012/10/01/2012;123(1):64–67. 10.1016/j.bandl.2012.07.006

54. Kato T. Principle and technique of NIRS-Imaging for human brain FORCE: fast-oxygen response in capillary event. International Congress Series. 2004/08/01/2004;1270:85–90. 10.1016/j.ics.2004.05.052

55. Chen Y, Chen Z, Estrada H, et al. Decoding paradoxical BOLD responses to transcranial ultrasound stimulation with concurrent optoacoustic magnetic resonance imaging. Sci Adv. Oct 2025;11(44):11. eadz1309. doi:10.1126/sciadv.adz1309

56. Glotzbach E, Mühlberger A, Gschwendtner K, Fallgatter AJ, Pauli P, Herrmann MJ. Prefrontal Brain Activation During Emotional Processing: A Functional Near Infrared Spectroscopy Study (fNIRS). Open Neuroimag J. 2011;5:33–9. doi:10.2174/1874440001105010033

57. Steber S, König N, Stephan F, Rossi S. Uncovering electrophysiological and vascular signatures of implicit emotional prosody. Sci Rep. Apr 2 2020;10(1):5807. doi:10.1038/s41598-020-62761-x

58. Herold F, Wiegel P, Scholkmann F, Müller NG. Applications of functional near-infrared spectroscopy (fNIRS) neuroimaging in exercise–cognition science: a systematic, methodology-focused review. Journal of clinical medicine. 2018;7(12):466.

59. Balaji S, Plonka N, Atkinson M, et al. Heart rate variability biofeedback in a global study of the most common coherence frequencies and the impact of emotional states. Sci Rep. 2025/01/25 2025;15(1):3241. doi:10.1038/s41598-025-87729-7

60. Chung S, Hwang H, Kim H, Hong J, Kim S, Han D. Changes in heart rate variability and hemodynamics of adolescents within the frontal cortex in response to face emotional stimulation. PLoS One. 2025;20(7):12. e0326204. doi:10.1371/journal.pone.0326204

61. Cowen A, Sauter D, Tracy J, Keltner D. Mapping the Passions: Toward a High-Dimensional Taxonomy of Emotional Experience and Expression. Psychol Sci Public Interest. Jul 2019;20(1):69–90. doi:10.1177/1529100619850176

62. Lee M, Choi H, Jo Y. Targeting emotion dysregulation in depression: an intervention mapping protocol augmented by participatory action research. BMC Psychiatry. Sep 2024;24(1):18. 595. doi:10.1186/s12888-024-06045-y

63. Ma H, Cai M, Wang H. Emotional Blunting in Patients With Major Depressive Disorder: A Brief Non-systematic Review of Current Research. Review. Front Psychiatry. Dec 2021;12:6. 792960. doi:10.3389/fpsyt.2021.792960

