## Supplementary Materials for "Auditory-induced Affective State Modulates Left DLPFC Engagement in Response to iTBS: An fNIRS Study in Healthy People"

**Supplementary Material**

**Section A. Target-Site Specificity of Affective State Modulation**

To test whether the affective state modulation of the HbO response to iTBS was target-site specific, a 3-way RM ANOVA was conducted with State (control vs. affective), Time Phase (during-stimulation vs. post-stimulation), and Site (left DLPFC vs. right DLPFC) as within-subject factors. A significant State × Site interaction emerged (F_1,35_ = 5.05, p = 0.031, partial η² = 0.126), indicating that the affective state differentially influenced HbO responses between the stimulated and contralateral hemispheres (Figure S2b). The State × Time Phase × Site interaction was not significant (F_1, 35_ = 3.13, p = 0.086, partial η² = 0.086), suggesting that this target site-specific modulation was broadly stable across both time phases.

Follow-up 2-way RM ANOVA within the right DLPFC revealed neither a significant main effect of State (F_1, 35_ = 0.72, p = 0.403, partial η² = 0.020) nor a State × Time Phase interaction (F_1, 35_ = 0.11, p = 0.740, partial η² = 0.003). Collectively, these results confirm that the affective modulation of iTBS-evoked HbO was localized to the stimulated DLPFC and absent in the contralateral hemisphere, arguing against a generalized affect-driven or systemic physiological artifact.

**Section B. Hemodynamic Feature Analysis Across Valence Polarities**

Extending our findings on the impact of auditory-induced affective states, we further compared the two valence polarities using quantitative metrics that capture complementary information to interpret the temporal and morphological characteristics of the hemodynamic response (Figure S1)^1^. Specifically, peak amplitude was used to measure response strength, time-to-peak (TTP) to assess latency from condition onset, and full width at half maximum (FWHM) to quantify response duration at half-peak amplitude^1^.

Regarding HbO, both conditions were characterized predominantly by a notable decrease. In the Positive condition, the peak amplitude reached -0.201 μM with a TTP of 64.6 s and an FWHM of 59.9 s, whereas in the Negative condition, the peak amplitude was -0.147 μM with a TTP of 70.6 s and an FWHM of 49.7 s. Differences between the two valence polarities were more pronounced in HbR: the Negative condition exhibited a substantially higher peak amplitude (0.409 μM vs. 0.174 μM), a shorter TTP (63.3 s vs. 74.8 s), and a narrower FWHM (73.2 s vs. 109.3 s) than the Positive condition. Thus, for HbR, the Negative condition elicited a faster and stronger peak response compared to the Positive condition, which produced a slower but more sustained response to iTBS, persisting into the post-stimulation phase.

From a developmental perspective, a previous infant study found that viewing happy faces elicited a gradual HbO increase, in contrast to the rapid peak and swift post-stimulus decline triggered by angry faces^2^. Although a similar feature was observed in a different chromophore (HbR) here, such temporal divergence may stem from an evolutionary adaptation: negative stimuli serve as threat signals requiring rapid avoidance, whereas positive cues facilitate approach-oriented communicative signals that necessitate more sustained neural processing^2^. For clinical translation, and given our finding that positive valence significantly reduced iTBS-induced pain, future work should investigate which valence polarity optimizes the overall treatment experience without compromising clinical effectiveness, if affective state-dependent iTBS proves therapeutically advantageous.

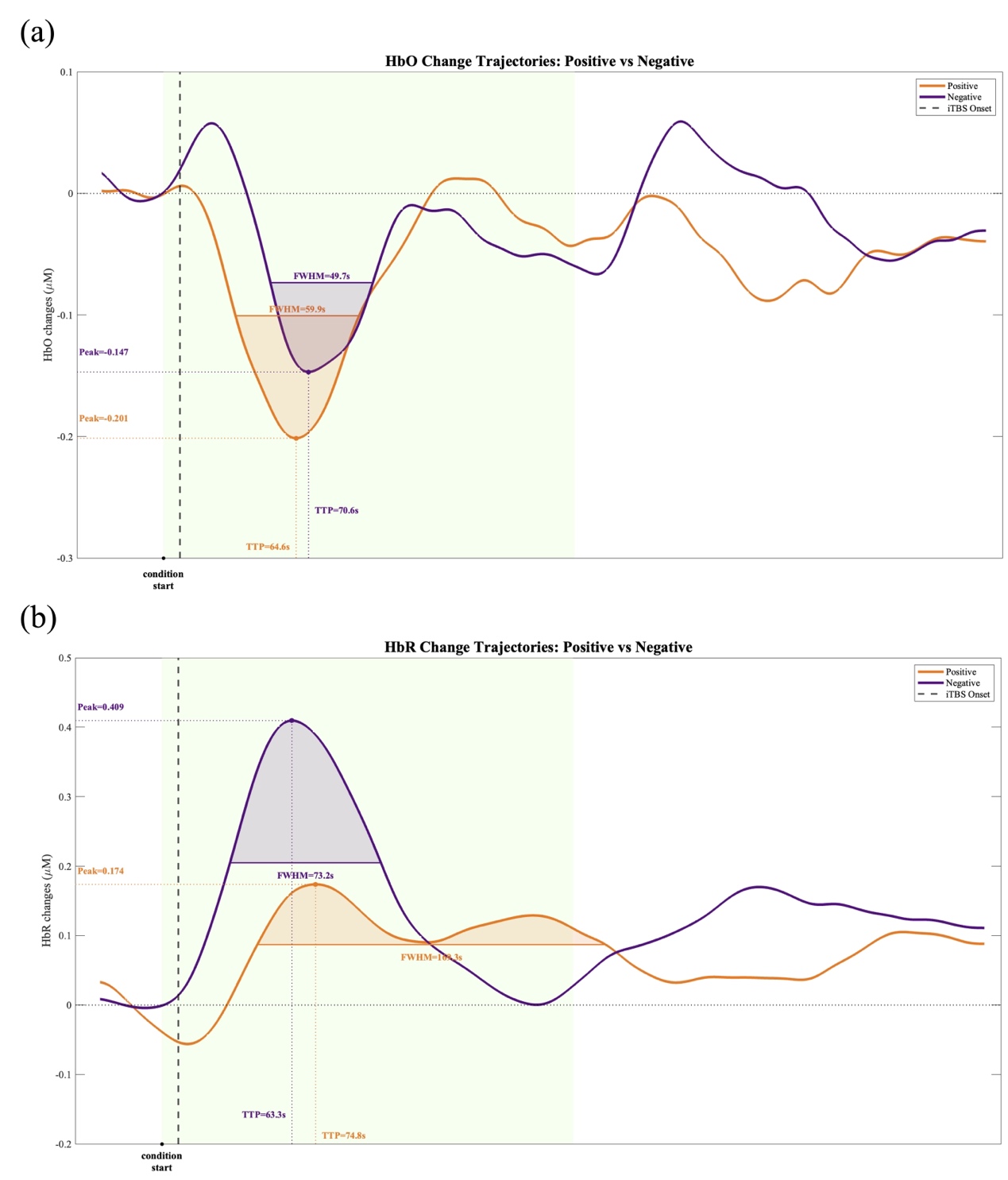
**Figure S1. Quantitative metrics characterizing hemodynamic changes in the Positive and Negative conditions.** Orange and purple solid lines represent the mean hemodynamic change trajectories over time for the Positive and Negative conditions, respectively. The green-shaded area indicates the 200 s during-stimulation phase. Marked features include peak amplitude, time-to-peak (TTP), and full width at half maximum (FWHM). **(a)** HbO; **(b)** HbR.

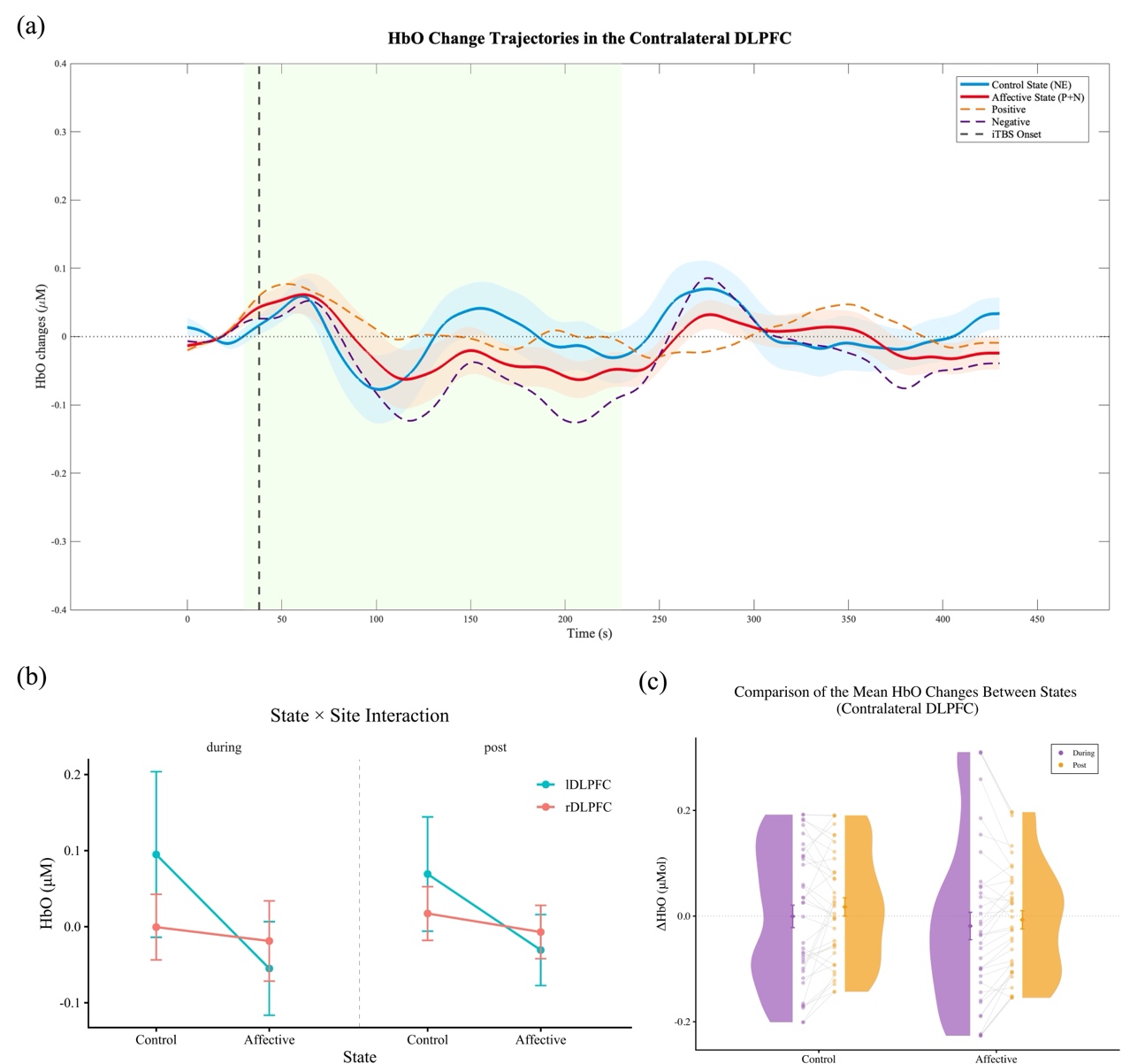

**Figure S2. Hemodynamic responses in the contralateral DLPFC and cross-hemispheric comparison.** **(a)** Real-time trajectories of HbO changes in the right DLPFC. **(b)** Mean ΔHbO in the left and right DLPFC across control and affective states during and post-stimulation. The State × Site interaction reached statistical significance. Error bars denote 95% confidence intervals. **(c)** Mean ΔHbO comparisons between control and affective states during and post-stimulation in the right DLPFC.

**Reference**

1.	Althobaiti M. Sensitivity analysis of the balloon model parameters in functional near-infrared spectroscopy simulation. *Journal of Neuroscience Methods*. 2025/12/01/ 2025;424:110599. doi:<https://doi.org/10.1016/j.jneumeth.2025.110599>

2.	Nakato E, Otsuka Y, Kanazawa S, Yamaguchi MK, Kakigi R. Distinct differences in the pattern of hemodynamic response to happy and angry facial expressions in infants — A near-infrared spectroscopic study. *Neuroimage*. 2011/01/15/ 2011;54(2):1600–1606. doi:<https://doi.org/10.1016/j.neuroimage.2010.09.021>
